# How to Develop Patient Centred Consulting during Workplace Learning in Postgraduate Medical Education? Opening the Black Box Using the Framework of Four Narrative Profiles for Consultation Performance

**DOI:** 10.1101/2025.01.14.25320523

**Authors:** Angelique Timmerman, Teresa Pawlikowska, Cees van der Vleuten, Jean Muris

**Affiliations:** Maastricht University, Department of Family Medicine, Care and Public Health Research Institute, Faculty of Health, Medicine and Life Sciences, Maastricht, the Netherlands; Royal College for the Surgeons of Ireland, University of Medicine and Health Sciences, Health Professions Education Centre, Dublin Ireland; Maastricht University, Department of Educational Development and Research, School of Health Professions Education, Faculty of Health, Medicine and Life Sciences, Maastricht, the Netherlands

**Author notes:** **Corresponding author:** Angelique Timmerman, Maastricht University, Department of Family Medicine, Faculty of Health, Medicine and Life Sciences, P.O. Box 616, 6200 MD Maastricht, the Netherlands, email address (AT).

## Abstract

**Context:** Four narrative profiles were previously developed as an evidence-informed framework for reflection and feedback on consultation performance in medical education. The profiles are grounded in four typologies mapped onto a conceptual framework, using the dimensions of doctor patient interaction (DPI) and medical expertise (ME) to classify overall performance.

**Objective:** Content validation of the narrative profiles derived from routine clinical consultations to inform a developmental roadmap for patient-centred consulting.

**Methods:** A qualitative study was performed in Family Medicine (FM) residency training, in which 11 first year and 7 third year FM trainees participated. The same FM assessor (n=11) observed a series of encounters of the trainee every three months during their training year, after which recurrent behaviours were described in a feedback report and overall consultation performance classified in one of the four typologies. Feedback reports (n=56) were categorised for each typology, coded on recurrent behaviours and then compared with the concordant narrative profile content.

**Results:** We identified overlapping recurrent behaviours and communication themes between the narrative profiles and feedback reports. For typology 1 (DPI+, ME-), the communication approach shows natural alignment with the patient, more exploration of patient concerns is needed, and the treatment plan needs connection to the reasons for consulting. In typology 2, active listening provides room for patient experience, yet patient centredness risks losing focus, while exploring of reasons for consulting is present. Inadequate responses to patient cues and losing structure in the consultation for typology 3, result in not grounding reassurance in clinical findings. For typology 4, curiosity towards patient’s concerns, may support understanding of symptoms and expectations.

**Conclusion:** A developmental road map for patient centred consulting was outlined, covering balancing medical tasks with exploring patient cues, enacting leadership in agenda management and integrating medical expertise in applied communication, providing focus and language for individual learning trajectories.

## Introduction

Communication is a core competency of physicians and has been positively associated with patient outcomes such as adherence to treatment recommendations and improved health and personal well-being and satisfaction (1) (2). Although communication is present throughout our daily lives, medical communication requires training and has become an important component of medical curricula (3) (4). The goals of medical communication have been clearly defined in literature (5) (6), however the application in a patient centred and contextualised manner during clinical encounters is less straightforward (7) (8).

Confronting learners with their communication in clinical encounters encourages them to reflect on consultation behaviours that need improvement and to practice alternatives (9) (10). Therefore, it is essential that learners receive regular and tailored feedback on their behaviours to support self-monitoring of consultation performance (11) (12). Feedback should be meaningful, authentic, and acceptable to promote a growth-oriented approach towards learning (13) (14). Current assessment methods often consist of standardised checklists that reduce communication to skills that must be routinely performed in every clinical encounter while communication is inherently contextualised and goal-directed (7) (15). The feedback provided may not be recognised as related to actual performance and therefore be dismissed by the learner as not credible or useful for learning (16) (17). Assessment practices that describe recurrent behaviours over multiple consultations, consider contextual influences and different levels of complexity of clinical cases and therefore can provide a more meaningful and realistic picture of learners’ performance.

In response to this identified educational need, we developed four narrative profiles in a Family Medicine (FM) residency training programme that describe recurrent behaviours and learning challenges for developing contextual adaptation (18). These narrative profiles are grounded in four typologies mapped onto a conceptual framework using the dimensions of doctor patient interaction (DPI) and medical expertise (ME); exemplary performance in these competences enables performing shared decision making (SDM) in the consultation. Given our goal of supporting learners in becoming proficient in their consultation performance, narrative profiles should balance sufficiently holistic descriptions of behaviours while providing the necessary detail to facilitate alignment of assessment and learning practices (19) (20). Their content is based on descriptions of recurrent behaviours observed by clinical supervisors observing digital consultations representing each of the typologies (18). Content validation of narrative profiles using real-life clinical encounters of multiple trainees with diverse medical problems and patient factors is important to support their transferability to educational practice.

The aim of this study is to conduct a content validation of the narrative profiles by comparing them with feedback from clinician expert assessors based on observed consultation performance in routine practice consultations of trainees. The following research questions are successfully addressed: How does feedback on trainee consultation performance, based on a series of real-life clinical encounters, relate to the recurrent behaviours of the narrative profiles? Can we discern recurrent patterns of recurrent behaviours from this comparison that outline an approach to develop patient centred consulting in medical education?

## Materials and methods

### 2.1. Study setting

This study was conducted in a three-year FM residency training programme in Maastricht, the Netherlands. During workplace learning, trainees develop their clinical competences in their routine encounters with patients in primary care. In the first and third year of training, the FM trainee works four days in a teaching practice under guidance of a FM supervisor, supplemented by a weekly educational release day at the training institution. During the second year the trainee participates in internships in other medical settings (elderly care, mental health, emergency care).

Formative assessment takes place both in FM practice and on the weekly release days, through observation and feedback on digitally recorded consultations, by supervisors, peer trainees and communication trainers respectively. Additionally, a separate evaluation of consultation performance is conducted by the training institution, for which the trainee must submit six self-selected video consultations, both in years 1 and 3, for assessment and feedback from an independent trained FM assessor.

### 2.2 Participants

Two groups of participants were involved in this study, consisting of 11 first year and 7 third year FM trainees, who followed the regular FM training programme and their internships in FM practice. Second, expert FM assessors (n=11), trained by the FM training institute to assess the regular consultation test in trainees, observed and provided feedback on the series of clinical encounters of these trainees. The assessor could be a FM supervisor with training in the assessment of consultation performance, but they did not supervise the specific trainee assessed during this study. The same FM assessor observed a trainee four times during the training year but was not involved in the regular training or familiar with the respective trainee.

### 2.3 Design and data collection

This qualitative study took place in parallel to the regular training trajectory of participating trainees, during a full training year. The study approach is outlined in a flow chart, summarising the consecutive stages of data collection and analysis (see Fig 1).

**Fig 1.**
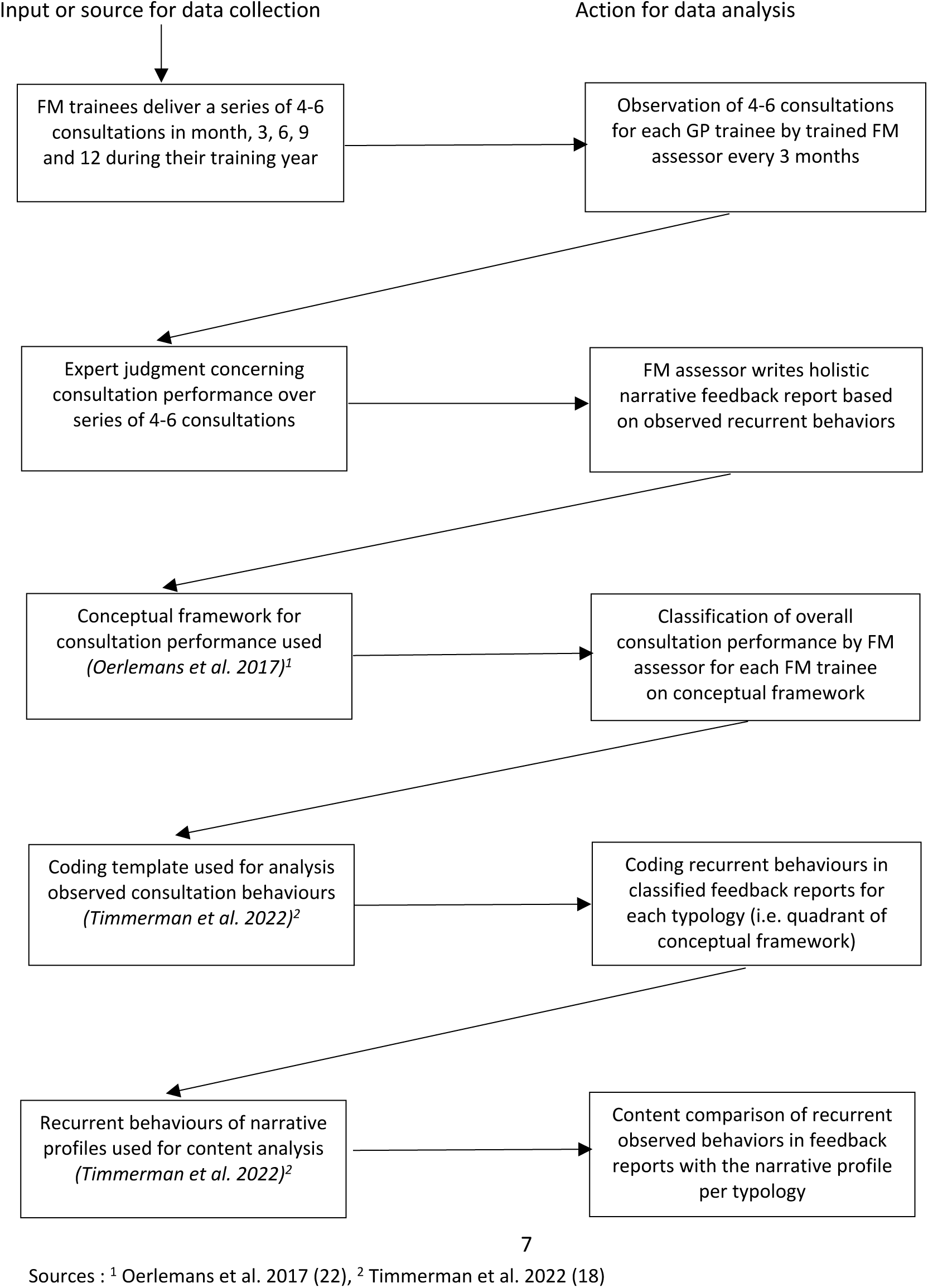
Flow chart of study procedure – data collection and analysis.

The design consisted of two related stages of data collection, repeated every three months: firstly, trainees were requested to record a series of routine practice consultations, in months 3, 6, 9 and 12 of their training year. This is intended to ensure the availability of consultations with a sufficiently diverse range of medical complaints and diagnoses according to the ICPC (International Classification of Primary Care) criteria to provide an overview of a trainee’s consultation performance (21). Secondly, FM assessors observed the recordings of trainees’ consultations uploaded in the secured digital assessment platform of the FM training institute (codific.com).

The focus was on describing recurrent behaviours over the 4-6 observed consultations for each trainee and formulation of personalised learning challenges. The assessors were instructed to make an overall judgment over the series of observed consultations on the conceptual framework for consultation performance, which maps the four typologies, with the dimensions of medical expertise and doctor patient interaction representing the horizontal (ME) and vertical (DPI) axes (see Fig 2).

**Fig 2.**
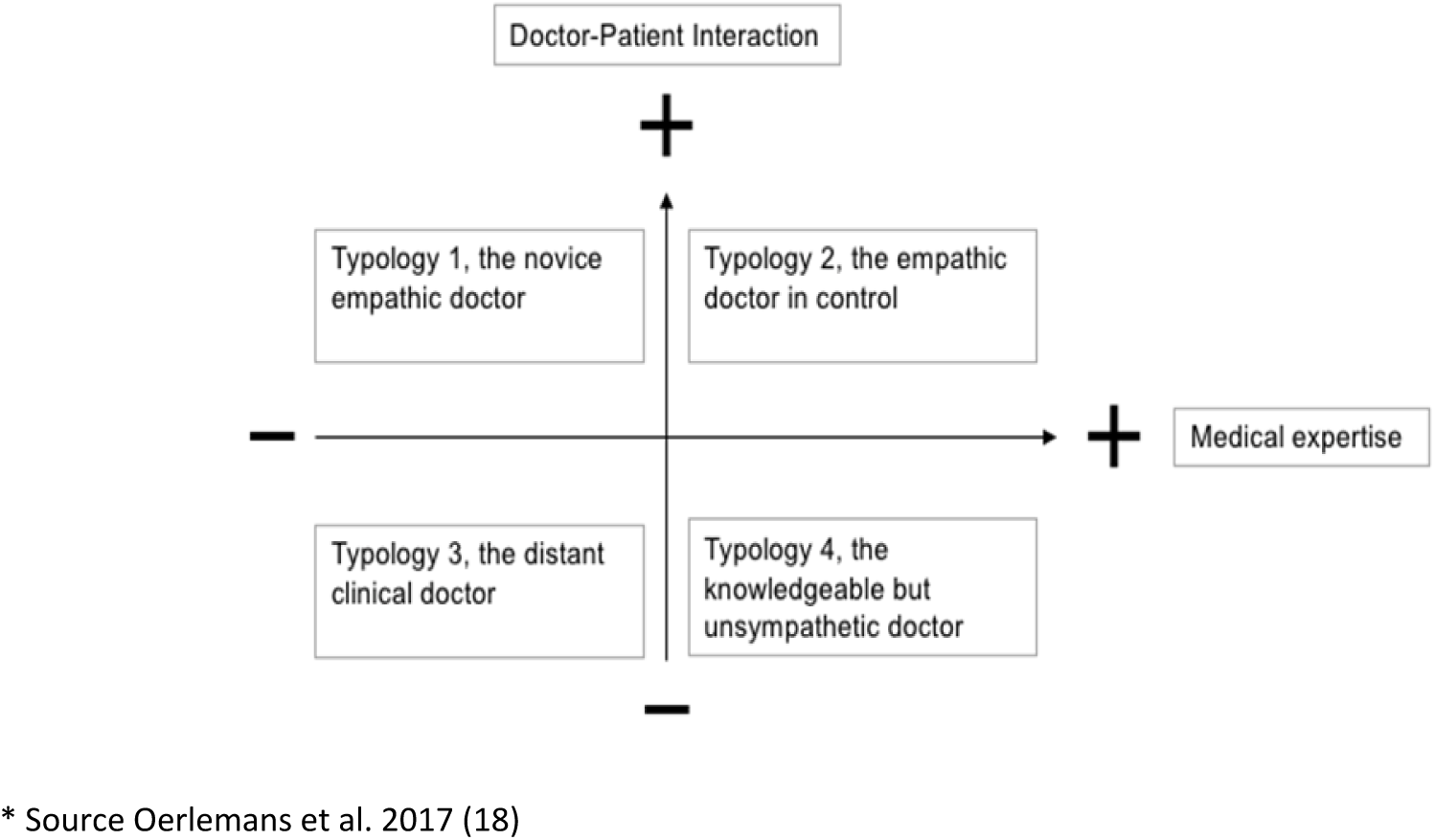
Conceptual framework for classifying recurrent behaviours in consultation performance using four typologies^*^.

The four typologies are: 1. Novice empathic doctor (DPI+, ME-), 2. Doctor in control (DPI+, ME+), 3. Distant clinical doctor (DPI-, ME-), 4. Unsympathetic but knowledgeable doctor (DPI-, ME+) (22). The recurrent behaviours representing each typology have been described in the four narrative profiles developed in our previous study (18).

In this study, we provided the FM assessors only with descriptions of consultation tasks related to doctor-patient interaction: building a relationship, structuring the consultation and communication; and medical expertise: medical knowledge and contextual exploration, as guidance for the classification of the overall trainee performance over the series of encounters. This approach enables the content comparison of the recurrent behaviours outlined in the feedback reports with those in the narrative profiles. The observation of the series of consultations and classification of overall performance by the FM assessors was repeated every three months and the FM trainees received a narrative feedback report four times during their training year.

The full period of data recruitment and collection took place between 1^st^ of April 2018 and 29^th^ of February 2024.

### 2.4 Study analyses

The study methodology was informed by thematic qualitative analysis to identify, analyse and assemble repeated behavioural patterns in the data (23). Data analysis was performed in two stages. Using the classification of overall trainee performance on the conceptual framework, each feedback report of the assessor over the series of observed consultations was grouped into one the four typologies. Followed by coding of the recurrent behaviours in the feedback reports and aggregating the identified codes for each typology. A coding template developed in a previous study was used, using the sensitising concepts of doctor patient interaction, medical expertise and shared decision making to code observed behaviours (18). The recurrent behaviours of the feedback reports were coded as either desirable or undesirable based on the reasoning of the FM assessor. Each feedback report was individually coded by two researchers and codes allocated were compared and discussed where inconsistencies in coding arose until consensus was reached.

In addition, a content comparison was performed between the codes identified over the series of feedback reports per typology and the recurrent behaviours of corresponding narrative profile. Similar and differing recurrent behaviours that concern the same consultation tasks (e.g. exploring patient context) of the narrative profiles and feedback reports were identified and assembled based on content analysis (24). As a research team we discussed these findings and reflected on recurrent consultation behaviours that were distinctive for each typology.

The distinctive behaviours were summarised in four diagrams representing each typology. We also identified resulting learning challenges to outline a roadmap for developing patient centred consulting in medical education.

### 2.5 Ethics

Ethics approval was obtained from the Ethical Review Board of the Netherlands Association for Medical Education (NMVO-ERB file number 597). Written informed consent was obtained from patients, trainees, supervisors and expert assessors before participation. We anonymised the verbatim transcriptions of the feedback reports.

## Results

### 3.1 Study characteristics

A total of 56 feedback reports were included: of these, 33 were from year 1 and 23 from year 3 trainees (see Table 1). The overall classification of trainee’s consultation performance for each series of 4-6 clinical encounters that informed the narrative feedback. This was 9 for typology 1 (DPI+, ME-), 33 for typology 2 (DPI+, ME+), 5 for typology 3 (DPI-, ME-), and 9 for typology 4 (DPI-, ME+). Across training years, it is noteworthy that all feedback reports in typology 1 were from year 1 trainees, and 8 out of 9 in typology 4 were from year 3 trainees.

**Table 1.**
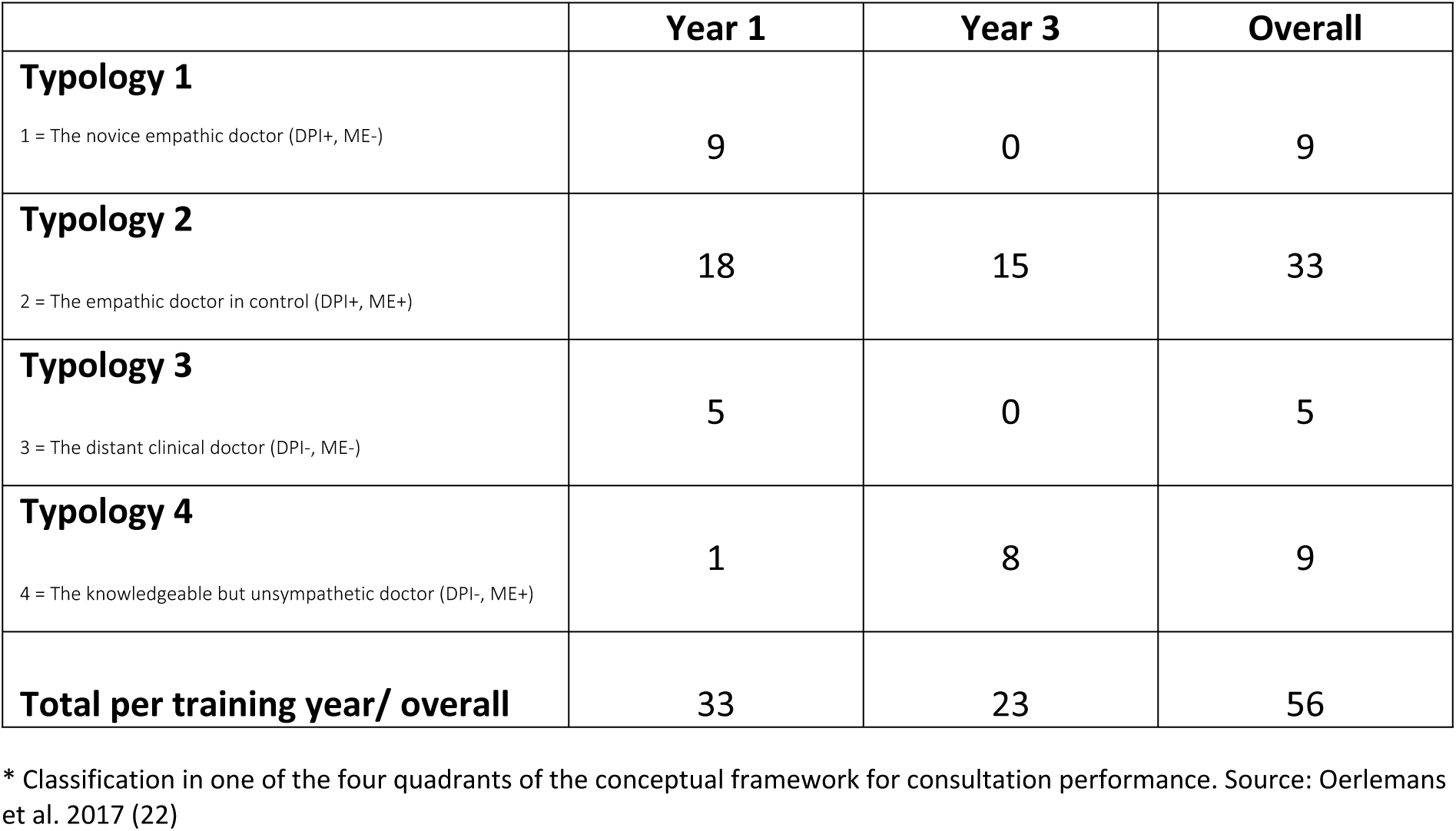
Overview of the number of classified feedback reports for each typology for consultation performance, per training year and overall^*^.

### 3.2 Study findings

In this section, we compare the recurrent behaviours of the four narrative profiles and the series of feedback reports as grouped per typology (see S1_Tables A-D). The behaviours in the narrative profiles and feedback reports are aligned in content and language in four diagrams. The diagrams show the comparisons between recurrent behaviours of the profiles and grouped feedback reports. Each diagram covers one quadrant (i.e. typology) of the underlying conceptual framework for consultation performance. The overlap in the diagram shows areas or characteristics of alignment in communication behaviours. In the non-overlapping left (i.e. narrative profile) and right (i.e. feedback reports) parts of the diagram behaviours are listed that share the same communication theme but are different in nature for both data sources. Recurrent behaviours identified from the feedback reports, yet not described in the narrative profiles are outlined, before describing the learning challenges arising from the content comparison of both data sources.

#### 3.2.1 Content comparison for typology 1 (ME-, DPI+): novice empathic doctor

This doctor aligns with the patient’s language and manners, shows some attention to the doctor-patient relationship, yet does not consistently explore patient concerns and the treatment regimen needs to be tailored more to the patient’s reasons for consulting. These recurrent behaviours of the first narrative profile are congruent with those in the feedback reports grouped in typology 1: the doctor shows a natural communication approach, does not sufficiently explore the patient’s concerns and feelings and a treatment plan tailored to the reason for consulting is lacking (see Fig 3A). Similar communication themes in the first narrative profile and feedback reports are agenda setting and providing information, but the behaviours described are slightly divergent in nature.

**Fig 3A.**
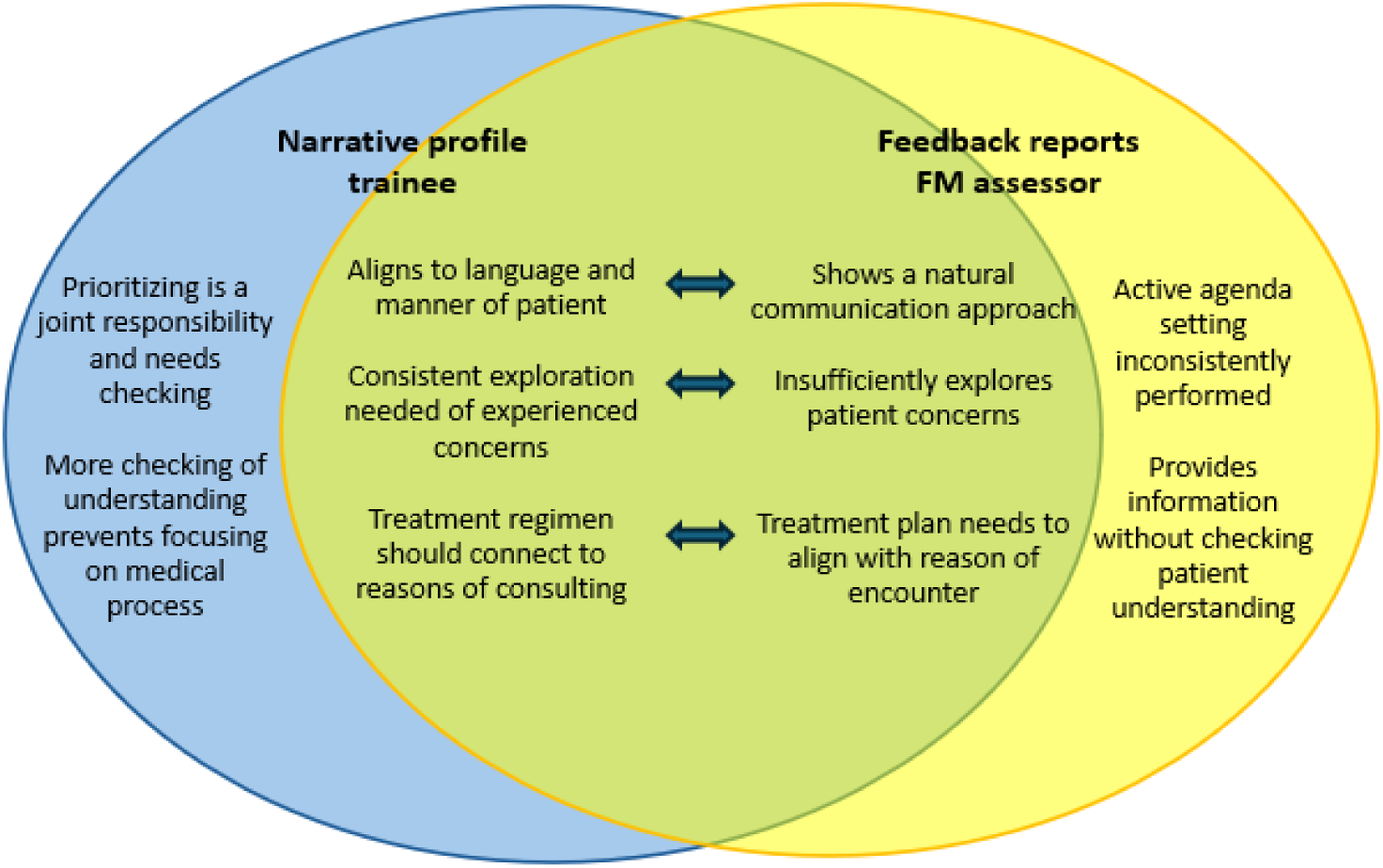
Diagram content comparison typology 1 (ME-, DPI+)

Recurrent behaviours emerging only from the feedback reports are that the doctor does not ask for other reasons of encounter and more explicit instructions are needed for the physical examination. The diagnosis and treatment regimen are adequate according to clinical standards and the doctor uses digital data sources to support explanations or provide additional information.

The learning challenge for this doctor is to develop confidence in acquired medical knowledge to free up mental space for exploration of patient concerns, in addition to performing a shared agenda setting to align the medical process with these concerns.

#### 3.2.2 Content comparison for typology 2 (ME+, DPI+): empathic doctor in control

The doctor has a curious attitude and the patient’s frame of reference is central during the clinical encounter, while active listening supports exploring the reasons for consulting. There is a risk of losing focus and overview when the emphasis on patient centeredness becomes too strong (see Fig 3B). Consistent with the recurrent behaviours in the feedback reports grouped in typology 2, this doctor connects to the patient’s experience by active listening and exploring the reason for consulting. In leaving much room for patient concerns, there is a risk of less structuring occurring during the encounter. Similar communication themes in the second narrative profile and feedback reports are: providing room to patients and integrating contextual factors into SDM, with slightly divergent behaviours.

**Fig 3B.**
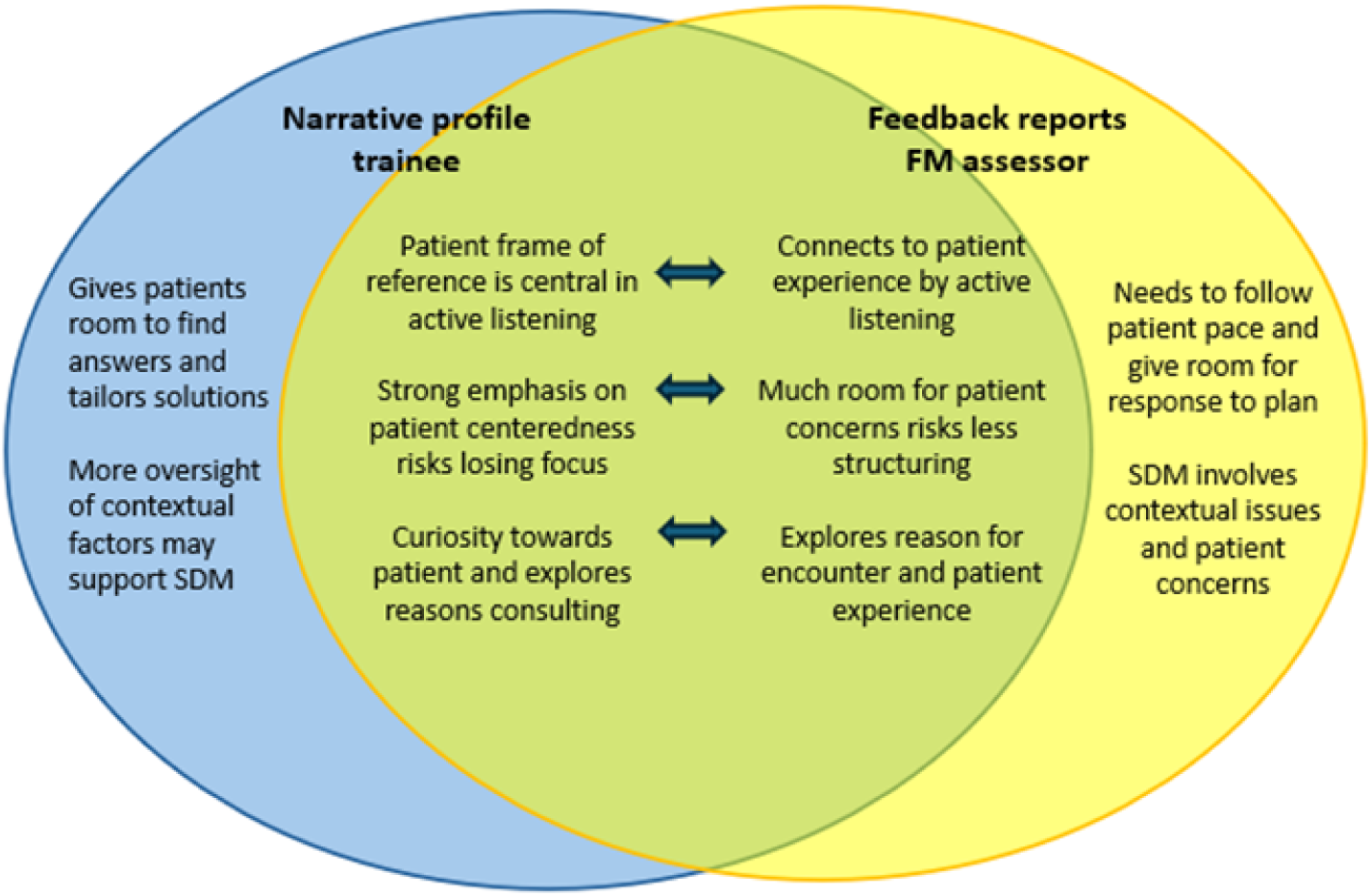
Diagram content comparison typology 2 (ME+, DPI+)

Recurrent behaviours that emerge only from the feedback reports are that the doctor does not ask key questions in the history taking phase and the physical examination should be more systematic. Explanation of diagnostic considerations and treatment regimen are clear and medically correct. Decision making can be optimised by including medical findings and not leaving the decision too much with the patient.

For this doctor, the learning challenge is to balance patient centeredness with performing medical tasks, through active agenda setting and integrate patient and doctor perspectives in SDM.

#### 3.2.3 Content comparison for typology 3 (ME-, DPI-): distant clinical doctor

The doctor does not respond adequately to patient cues and empathy is lacking while limited medical knowledge leads to attempts to control the dialogue and consultation flow. Reassurance of the patient becomes more effective if information is based on clinical findings. The feedback reports grouped in typology 3 confirm these recurrent behaviours in that the doctor often ignores information or requests from the patient during the encounter and a lack of medical knowledge causes a chaotic explanation phase. To prevent patient worries, diagnostic hypotheses shared with the patient should be grounded in a physical examination (see Fig 3C). Similar communication themes in the third narrative profile and feedback reports are structuring the consultation and clarifying the nature of symptoms, albeit with slightly different behaviours described.

**Fig 3C.**
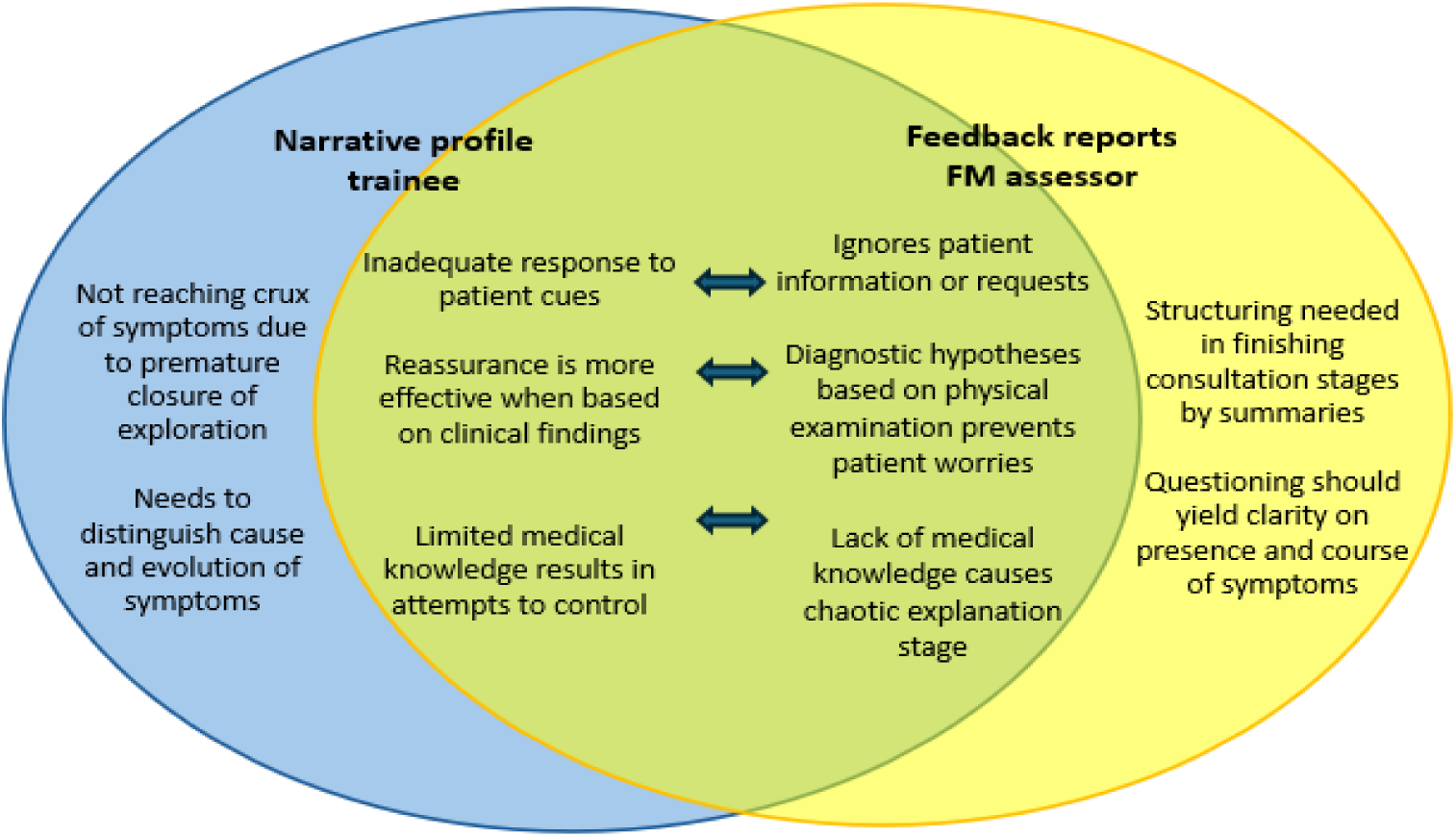
Diagram content comparison typology 3 (ME-, DPI-)

Recurrent behaviours that emerged only from the feedback reports are that the doctor does not fully explore the patient’s needs, often leading to several initial complaints presented. The doctor asks questions about potential alarming symptoms. During decision making, the doctor gives alternative options only if the patient disagrees with the treatment regimen. Clinical encounters can generally be lengthy due to a lack of focus and pace.

The learning challenge for this doctor is to ground a diagnostic approach in medical knowledge and use structuring skills to meet the needs of the consultation stages, and to help navigate between exploring patient cues and addressing medical questions.

#### 3.2.4 Content comparison for typology 4 (ME+, DPI-): knowledgeable, unsympathetic doctor

In typology 4, the doctor’s focus is on the medical pathway using a diagnostic approach. It is necessary to connect with patient views to identify their expectations. Being curious about the patient may support connecting to what is needed in a relational way. From the feedback reports grouped in typology 4, a picture arises of a doctor who prefers to lead without being interrupted by patients expressing their concerns. There is limited exploration of the meaning of symptoms and concerns and curiosity towards the patient as a person is inconsistently present (see Fig 3D).

**Fig 3D.**
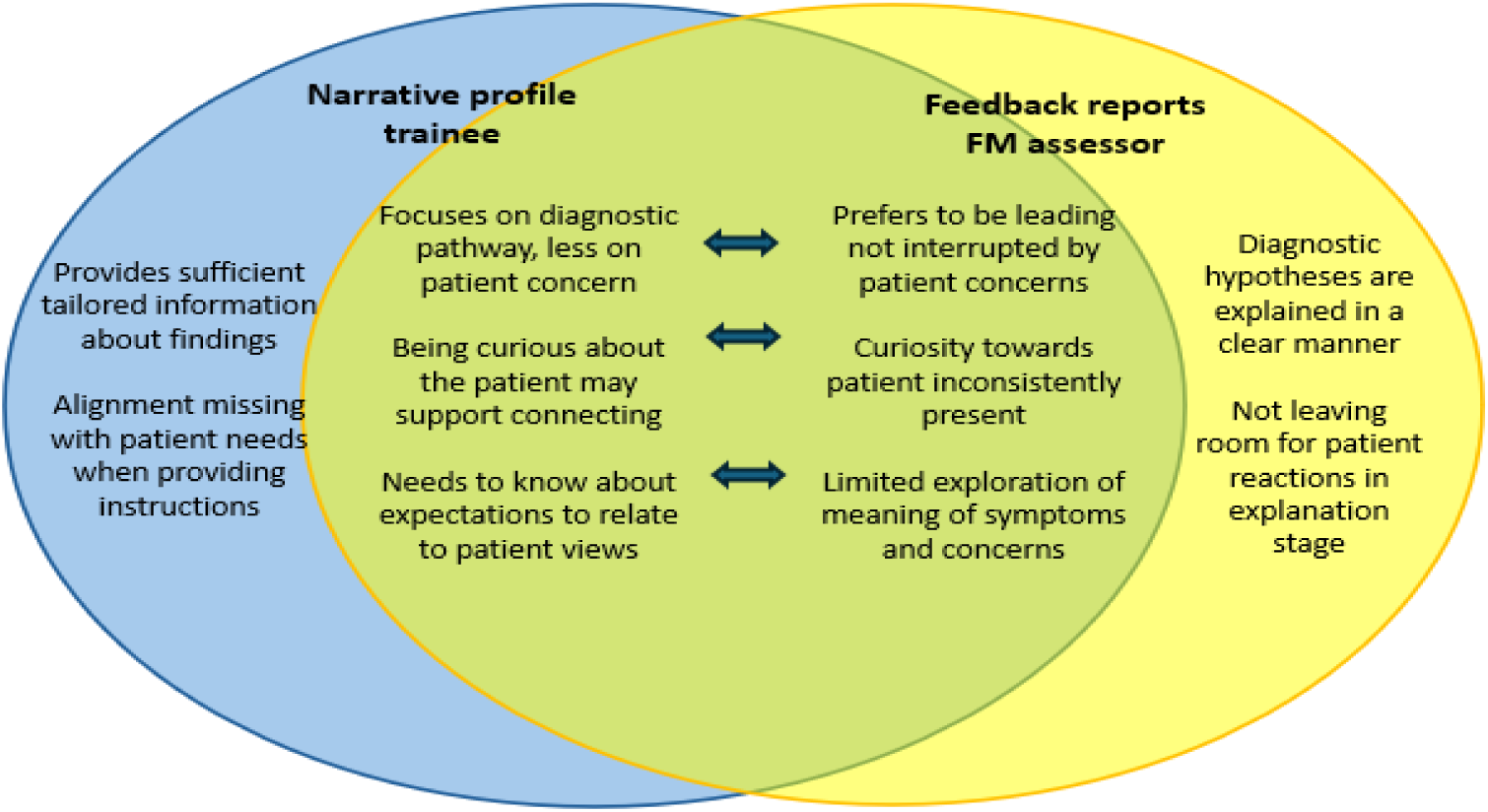
Diagram content comparison typology 4 (ME+, DPI-)

Similar communication themes in the fourth narrative profile and feedback reports are providing information and responding to patient needs. The described behaviours relate to the explanation and planning phase.

Recurrent behaviours that appear only in the feedback reports are that the doctor insufficiently reflects and explores the patient’s emotions and only states the reason for the encounter when expressed by the patient. Additional reasons for encounter are not explored, often resulting in a new complaint at closure that is addressed by the doctor without being explored.

For this doctor, the learning challenge is to leave room for a natural consultation flow and patient initiative, while fostering collaboration through exploring the meaning of (non)verbal patient cues and reaching decisions that include the patient’s agenda.

## Discussion and Conclusion

### 4.1 Discussion

The main aim of this study was to perform a content validation of four narrative profiles for consultation performance by comparing them with expert feedback on a series of real-life clinical encounters of trainees. The recurrent behaviours described in the feedback reports were overall similar compared to the content of the narrative profiles as we were able to identify both overlapping communication behaviours and themes. Recurrent behaviours in the communication themes were only slightly different in nature when the content of the narrative profile and feedback reports were contrasted. In addition, we could identify complementary behaviours from the feedback reports to enrich the content of each narrative profile. This content validation also resulted in themes that can operationalise an approach for developing patient centred consulting (25) (26) during medical education.

In this section we discuss how this approach can support learning for each of the typologies mapped on the conceptual framework for consultation performance, reflecting three themes: balancing performing medical tasks with exploring patient cues, and enacting leadership in prioritising clinical and patient needs, while integrating medical expertise is conditional for effective consultation performance. The first theme is *balancing medical tasks with exploring patient cues*. For typology 1 (ME-, DPI+) this involves exploring the patient’s concerns to formulate a treatment regimen that connects with the reasons for consulting. As in typology 2 (ME+, DPI+), keeping an overview of the patient’s concerns and clinical needs supports focus when performing medical tasks. In typology 3 (ME-, DPI-) grounding diagnostic hypotheses in clinical findings supports addressing patient needs and reassurance. In typology 4 (ME+, DPI-) this is reached through focusing less on the medical pathway and more on the patient’s concerns provides room to explore the experienced meaning of symptoms.

**Fig 4A.**
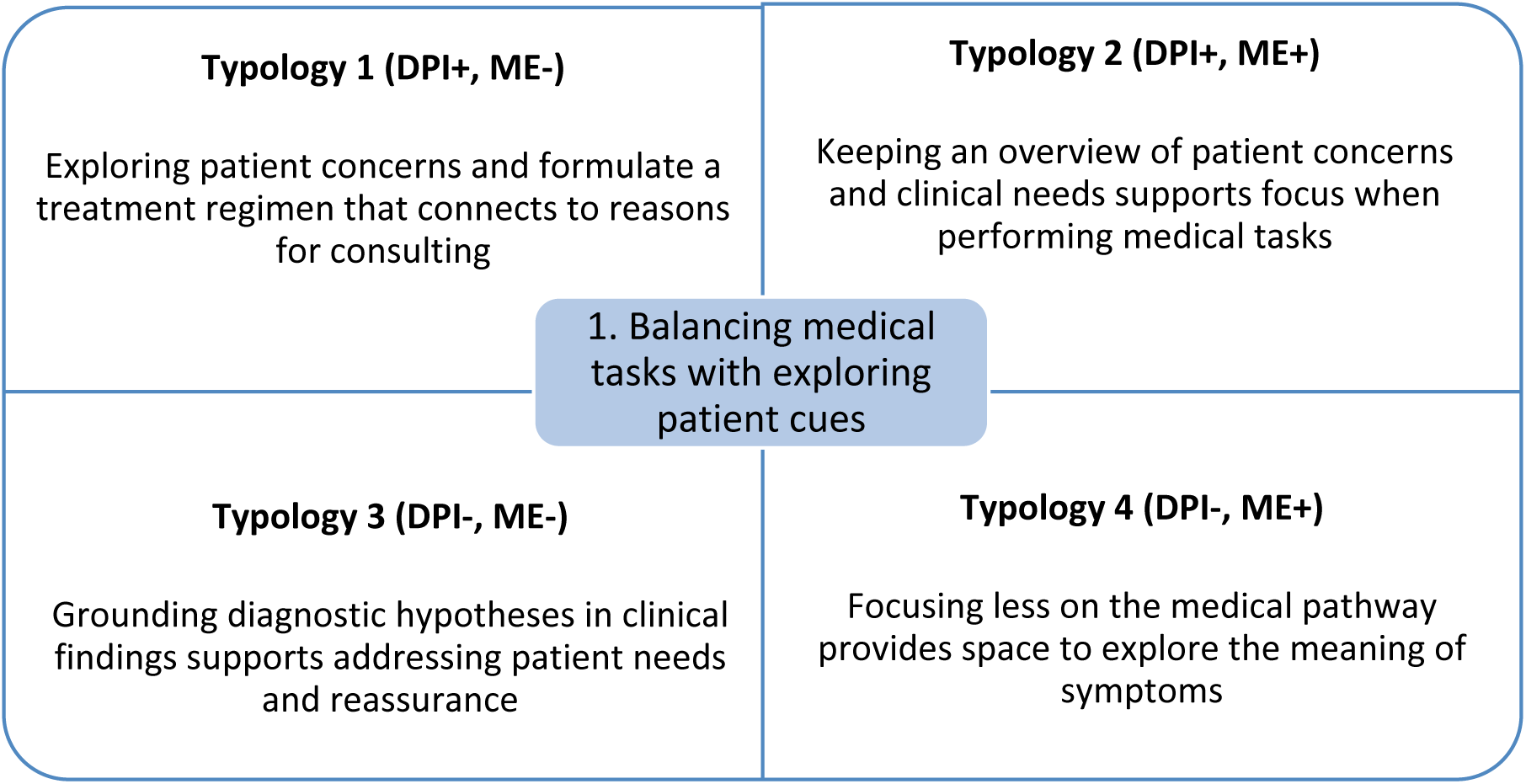
Developmental roadmap for patient-centered consulting using four typologies – theme 1.

Second is the theme of *enacting leadership in prioritising clinical and patient needs in the consultation*. In typology 1, shared agenda setting supports ascertaining of the patient’s needs and priorities, while in typology 2 active agenda setting can support more oversight of contextual factors to perform SDM. For typology 3, applying structuring skills during the consultation supports navigating between exploring patient cues and medical questions and in typology 4 providing room for patient initiative may result in decision making that includes the patient’s agenda.

**Fig 4B.**
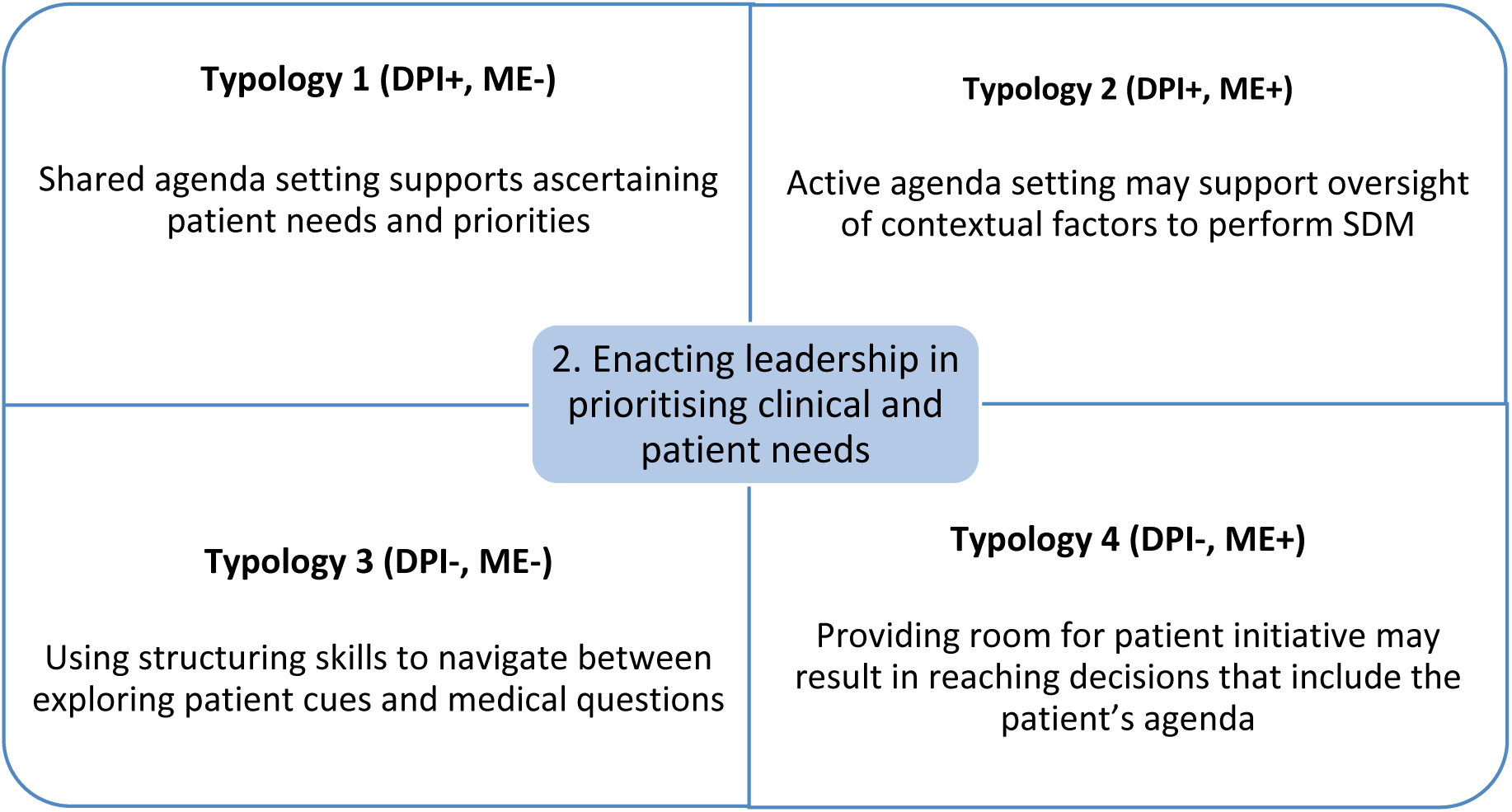
Developmental roadmap for patient-centered consulting using four typologies – theme 2.

A third theme, *integrating medical expertise with applied communication* is required for an effective consultation performance: developing confidence in medical expertise in typology 1, combining doctor and patient perspectives in SDM for typology 2, grounding a diagnostic approach in medical knowledge and in clinical reasoning for typology 3 and alignment of medical knowledge with patient needs in typology 4. communication applied still needs to be contextualised in each encounter, depending on the medical, patient and consultation factors present (29) (30).

**Fig 4C.**
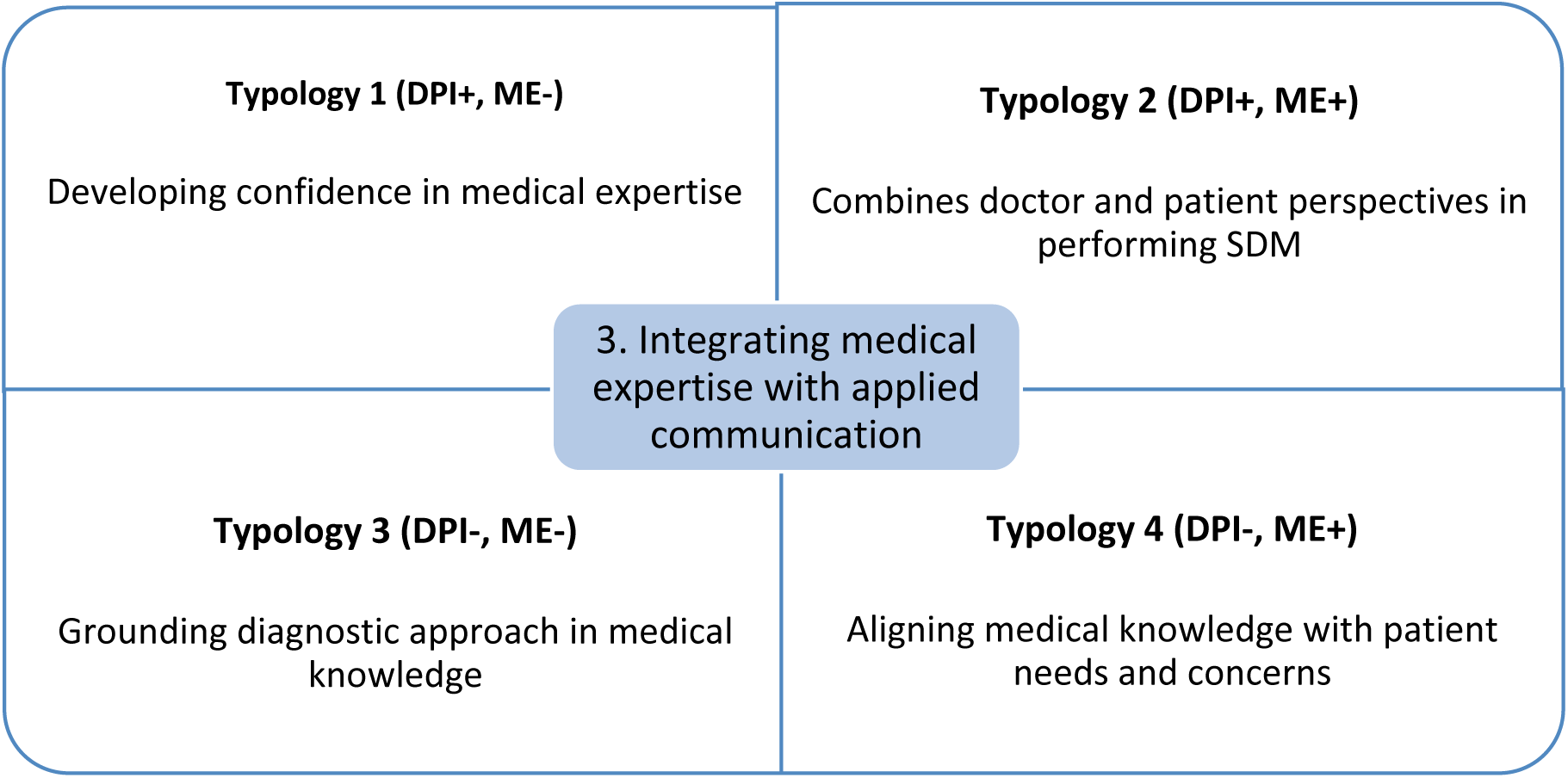
Developmental roadmap for patient-centered consulting using four typologies – theme 3.

These overall themes can be considered as learning challenges, to support the development of patient centred consultation skills. In doing so, we wanted to tie in with the ongoing development of the concept of patient centeredness, which can be narrowed down to three areas during medical consulting: understanding the patient’s illness and life experience, developing a doctor-patient relationship with shared guidance and coordinating care in the broader context of treatment (27) (28). Although these areas operationalise a participatory approach to clinical care, the In mapping the overall themes for each typology, we conceptualised a road map for patient centred consulting to develop the capacity to flexibly adapt communication during the clinical encounter (18) (22). We intend to support the process of learning communication during medical training which often takes place implicitly (31) in providing holistic anchors to facilitate reflection on actual consultation performance of learners. To develop contextual adaptation, learners and their educators should become sensitive to challenging patient cues that may stimulate reflection and feedback on applied communication during clinical encounters (32). Often triggers for learning are present in a less noticeable way and standards for what constitutes good consultation performance are hard to define given the complexity of the clinical encounter and interactive nature of communication (33) (34). That is why we do not only provide an evidence-informed framework for developing patient-centred consulting in medical education, but also language for feedback in signalling and supporting learners’ longitudinal progress.

#### 4.1.1 Strengths and limitations

The strength of the narrative profiles for consultation performance is their grounding in real-life clinical encounters of trainees. The recurrent behaviours described can be generalised over medical problems and contexts using a series of trainees’ clinical encounters in this study, which enriches our previous study, where we used one exemplary consultation for each typology during interviews with supervisors to develop the narrative profiles (18). The current study supports the transferability of the narrative profiles; for extrapolation the narrative profiles need to be applied in other medical training settings, as this study focused only on FM residency training (35).

The fact that trainees were predominantly classified in typology 2 (ME+, DPI+), indicated that they were already performing at a proficient level, followed by classification in typology 1 (ME-, DPI+) and typology 4 (ME+, DPI-). This performance is consistent with the level of postgraduate medical education, even though more trainees from year 1, rather than from year 3 participated, which is due to more focus on developing consultation skills early in the FM training programme.

In the content comparison between the narrative profiles and the feedback reports, we identified more complementary behaviours in typologies 3 (ME-, DPI-) and 2 (ME+, DPI+), located at the extremes of the learning spectrum: under and well performing trainees, which may indicate that formulating feedback is easier for these learners. This finding is in alignment with that supervisors in the earlier study were better able to identify under- and well performing trainees during observations of their consultation performance in clinical encounters (22). For the typologies 1 (ME-, DPI+) and 4 (ME+, DPI-), this may indicate that it is more difficult to signal and determine what intermediate performance is, which may compromise the content validation of the corresponding narrative profiles based on real-life clinical encounters of trainees.

A limitation is that our study design was labour intensive, which restricted the potential for dual observations of encounters and triangulation of expert perspectives. There is also a risk of stigmatisation by profiling learners’ progress, as their performance may still vary across consultations depending on the time available, the medical complaint presented and patient characteristics (12)(30). We, therefore, recommend further research to explore recurrent behaviours of learners whose performance was classified in one of the typologies over time and get insight into the longitudinal development of patient-centred consulting.

### 4.2 Conclusion

Narrative profiles for consultation performance compared favourably with expert feedback over observations of a series of clinical encounters of trainees. An approach to patient centred consulting has been outlined grounded in this content validation, which signposts a developmental road map for each typology of the conceptual framework. The following themes guide this approach: striking a balance between performing medical tasks with exploring patient cues, enacting leadership in prioritising clinical and patient needs during the consultation and integrating medical knowledge into applied communication. The learning challenges outlined for each typology can serve a dual purpose: providing a framework that provides focus in monitoring consultation performance and for feedback on learning practices of patient centred consulting.

### 4.3 Practice implications

The developmental road map described for each typology provides content focus and language to support learning dialogues across the continuum of medical education: what recurrent behaviours are observed in learners’ consultation performance, what skills are important to develop and what behaviours need to be practiced in clinical encounters? In this manner, educators and learners create a shared frame of reference while reflecting on medical knowledge and communication, to support identifying learning challenges for patient centred consulting that need to be addressed.

The developmental road map for patient centred consulting can also be used to inform narrative feedback from educators ‘in the moment’ during learning activities and foster performance improvement (36). Yet feedback ‘over time’ assists the learner in reflecting on learning progress and focusing on consultation behaviours that need to be practised. The narrative profiles can support this calibration through outlining consultation behaviours for each typology in more detail (37). The narrative profiles are not intended to be used as a prescribed guideline during medical education, but rather as a supportive tool for educator-learner interactions to provide focus and direction when developing learner action plans and co-creating learning activities for experimenting with alternative communication behaviours (14) (38). For the lifelong learning of physicians, developing feedback tools containing rich narrative information promises to encourage self-directed learning of patient centred consulting, and the narrative profiles can hopefully contribute to this purpose.

## Data Availability

All relevant data are within the manuscript and its Supporting Information files.

## Acknowledgments

We are grateful to the 18 trainees who participated in this study, their supervisors for enabling data collection and the 11 assessors for observing all consultations and writing the feedback reports. And a big thanks to Demi Schijlen and Ramona Guerrieri for their support in analysing all the feedback reports and the FM training institute for facilitating the recruitment of participants.

## Supporting information

S1 **Table A. Content comparison of narrative profiles and feedback reports of Family Medicine trainees – grouped for typology 1**

S1 **Table B. Content comparison of narrative profiles and feedback reports of Family Medicine trainees – grouped for typology 2**

S1 **Table C. Content comparison of narrative profiles and feedback reports of Family Medicine trainees – grouped for typology 3**

S1 **Table D. Content comparison of narrative profiles and feedback reports of Family Medicine trainees – grouped for typology 4**

